# Re-shaping professional boundaries to scale-up HIV pre-exposure prophylaxis (PrEP) services: collaborative care and power dynamics in Belgium

**DOI:** 10.64898/2026.07.14.26357825

**Authors:** Jef Vanhamel, Karina Kielmann, Thijs Reyniers, Gert Scheerder, Christiana Nöstlinger

**Author notes:** Corresponding author: Dr Jef Vanhamel, Postdoctoral Research Fellow, School of Public Health, Faculty of Health Sciences University of the Witwatersrand, 27 St Andrew’s Road, 2193 Parktown, Johannesburg, South Africa.

## Abstract

Scaling HIV pre-exposure prophylaxis (PrEP) services in health systems will require collaboration and clear role distinction among professionals, and between specialist and primary care. This study examined how power dynamics shape efforts to expand PrEP care beyond specialised HIV clinics in Belgium. We conducted semi-structured interviews with 36 HIV clinic providers and two community-based organisation (CBO) representatives, and 16 online group discussions with general practitioners (GPs). We analysed data thematically, guided by the concepts of collaborative and competitive power to examine how providers negotiated expertise and role division in PrEP delivery across professional and organisational boundaries. We found that reimbursement regulations anchored PrEP initiation and follow-up within HIV clinics, embedding specialist jurisdiction in care pathways. HIV specialists reinforced this position by drawing on their recognised expertise in HIV medicine to justify clinical coordination and authority in determining standards of care. GPs emphasised accessibility and preventive care roles but made limited claims to PrEP provision, linked to misaligned organisational incentives, role blurring, limited training opportunities, and the historical concentration of HIV care in specialist services. CBOs facilitated access, enabling coordination between vulnerable communities and clinics while remaining weakly embedded in formal care structures. Findings show that expanding integrated PrEP services beyond specialised care is not only shaped by operational issues such as training and resources but also by the structural dynamics of regulations, institutional mandates, and professional jurisdictions that influence collaboration. Effective scale-up will require policies that align incentives, clarify responsibilities, and support collaboration across specialised, primary care, and community settings.

**Highlights:**

- Collaborative pre-exposure prophylaxis (PrEP) delivery is shaped by power dynamics that operate differently across professional and organisational boundaries.
- Competitive power is embedded in regulation that favours specialist jurisdiction over PrEP delivery.
- Collaborative power develops where organisational and structural arrangements support shared-care, more readily available within HIV clinics.
- Interorganisational collaboration remains constrained by regulatory, organisational and historical factors.
- Scaling integrated PrEP services will require structural and institutional change, not only shifts in individual providers’ attitudes, motivation or (clinical) capacity.

## 1. Introduction

### 1.1 Background

More than a decade after HIV pre-exposure prophylaxis (PrEP) was first approved as a highly efficacious biomedical HIV prevention intervention, expanding its global roll-out remains a public health priority to reduce HIV incidence [1]. The question of how best to mobilise health system resources to effectively expand PrEP service delivery has sparked interest in scale-up strategies such as task-shifting and task sharing, integrated care, and de-centralised service delivery models involving community- or primary care-based PrEP provision [2–6]. Many of these strategies rely on the coordinated efforts of professionals from diverse disciplines and organisations. However, the dynamics underlying interprofessional and interorganisational collaboration within PrEP delivery remain poorly documented and understood.

Research on provider-related aspects of PrEP has primarily focused on individual-level behaviours and cognitive factors (e.g. provider knowledge, attitudes and willingness to prescribe PrEP) [7–9]. Few studies have explicitly examined the power dynamics and tensions resulting from shifts in role division and disruption of traditional disciplinary boundaries during PrEP scale-up. Yet conflicting views on the scope of practice and perceived accountabilities among providers in PrEP service delivery have been debated. For example, during the early roll-out of PrEP in the United States, researchers pointed to a “*purview paradox*” whereby neither HIV physicians nor primary care physicians viewed PrEP provision as part of their clinical responsibilities [7,10]. In Australia, HIV specialists expressed concerns that general practitioners (GPs) lacked the cultural competence and clinical expertise to deliver LGBTQI+ affirming care, prompting resistance to GP involvement during initial phases of the PrEP roll-out [11].

Questions of expertise and professional jurisdiction have been a longstanding and recurrent debate in the HIV response. This debate can be traced back to the evolution of HIV as a distinct clinical and professional discipline, with responsibilities around care and treatment traditionally being assumed by physicians, and health promotion and prevention being delegated to lower-level cadres such as nurses [12]. The increasing availability of HIV treatment amidst health workforce shortages has led to task-shifting away from physicians towards other professionals, especially in health systems with a high HIV burden [13,14]. The advent of PrEP has further contributed to the medicalisation of HIV prevention, blurring traditional professional boundaries between treatment and prevention, and fuelling calls to extend PrEP delivery to a broader range of professionals including nurses, pharmacists, GPs, community health workers and lay providers [15–18]. The creation of biomedical tools to manage risk and optimise prevention creates new fields of practice, in which professional ownership and roles are fragmented, forcing continuous re-negotiation of boundaries between specialists and generalists [19]. Debates over who should provide PrEP are often polarising, and improving collaboration may benefit from a better understanding of how provider roles and scopes of practice are negotiated within the systems where PrEP is delivered.

Belgium presents an interesting case to examine evolutions in the professional scope of practice around PrEP delivery. Since 2017, PrEP services are delivered via 12 specialised HIV clinics, which are generally located in secondary or tertiary health facilities. In addition to the fee-for-service remuneration of specialist physicians, these HIV clinics receive dedicated government funding to organise multidisciplinary care: services from nurses, dieticians, social workers, psychologists and sexologists are offered free of charge to patients enrolled in HIV care. Only infectious disease specialists operating in these HIV clinics are officially authorised to prescribe reimbursed PrEP [20]. However, in previous work, we have shown discrepancies between policy and practice, including, for example, the adoption of strategies like task-shifting to nurses and GP involvement to alleviate the burden on overstretched HIV clinics [21].

In Belgium, GP involvement in PrEP care has not received much policy attention, despite regional health authorities providing GPs a mandate to provide preventive services in primary care and GPs being responsible for diagnosing the majority of all annually reported new HIV cases [22,23]. Belgian GPs are predominantly reimbursed on a fee-for-service basis, while a minority works in a capitation system receiving a monthly fee [24]. Despite their neglected role in PrEP care, GPs in Belgium have shown increased interest in participating in PrEP delivery [25]. Since 2023, revision of the PrEP reimbursement regulations allows GP involvement in follow-up care, albeit only for two of the four annual visits, and, paradoxically, without granting them formal PrEP prescribing authority [26]. These developments reflect underexplored assumptions about what constitutes good-quality PrEP care, with important implications for resource allocation and organisation of PrEP services.

### 1.2 Theoretical framework

This paper draws on theoretical frameworks that explore professional collaboration in the context of intersecting roles. Specifically, we consider how providers perceive and implement collaboration in PrEP care while maintaining relative autonomy in their scope of practice. Two concepts are particularly useful here: competitive and collaborative power [27–30]. Competitive power, as described by Nugus *et al.*, is leveraged by clinicians from one professional domain or hierarchy to protect their boundaries from incursion by others. Collaborative power involves interdependent participation, shared decision-making and mutual accountability within teams [30]. The constant interplay between competitive and collaborative power means that defining and assuming roles and team structures across professional boundaries is not static, but rather a dynamic, context-dependent and continuously negotiated process [31,32]. Put differently, providers have agency over their actions and decisions while their capability to collaborate is also underpinned by systemic and structural determinants that inevitably involve power dynamics [31].

Interorganisational collaboration introduces additional complexity beyond intra-organisational team dynamics [33,34]. While providers within a single organisation share common accountability structures, resources and communication channels, collaboration across organisations requires bridging fundamentally different institutional contexts, each with their own mandates, funding mechanisms, and professional cultures [33]. Work from organisational sociology demonstrates that interorganisational collaboration depends on boundary-spanning activities – the work that actors do to create connections and coordinate action across organisational divides [35] – and on relational coordination, characterised by shared goals and knowledge, and mutual respect among actors in different organisations [32,36,37].

We make use of concepts of competitive and collaborative power to examine how healthcare providers in Belgium perceive their own role and the role of others in PrEP care, as well as how these power dynamics influence enactment of a collaborative approach to PrEP care at two levels: (1) among multidisciplinary teams within HIV clinics, and (2) across organisational boundaries, where HIV specialists, GPs and community-based organisations (CBOs) attempt to coordinate care. This dual-level analysis allows us to identify both shared dynamics and distinct challenges in fostering collaboration within and across organisational settings.

## 2. Methods

### 2.1 Study design

This study adopted an interpretivist qualitative design informed by critical social science perspectives on professional boundaries and power [38]. It was conducted as part of a larger interdisciplinary implementation research project aimed at informing the roll-out of PrEP in Belgium. We drew on data from two interlinked sub-studies: one involving semi-structured interviews with different HIV care providers in eight Belgian HIV clinics [21]; and another involving online group discussions with Belgian GPs [25]. Data were collected between November 2020 and May 2022, complemented by a review of two key policy documents governing the implementation of PrEP in Belgium [20,39].

### 2.2 Sampling and data collection

Details of sampling and data collection procedures for both studies are provided elsewhere [21,25]. Briefly, for Study 1, eight out of twelve invited HIV clinics across Belgium agreed to participate. During site visits we conducted interviews with purposively selected key informants and different types of PrEP providers. Key informants included senior physicians and staff with coordinating responsibilities in PrEP care, while PrEP providers included staff with critical roles in different steps of PrEP care. All those invited agreed to participate. We conducted 36 interviews (10 key informants and 26 care providers, including infectious disease physicians (9), nurses (10), psychologists/sexologists (4), and GPs attached to HIV clinics (2)). The number of interviews was primarily determined by purposive sampling across clinics rather than by data saturation, as interviews aimed to capture variation in roles and organisational context within and across clinics. We also conducted two additional interviews with CBO representatives – providing sexual health services to marginalised populations (e.g. undocumented sex workers) – who were identified as key collaborators of HIV clinics. All interviews were conducted in Dutch, French or English by a researcher with a medical background trained in qualitative research (JV) and lasted between 40 and 60 minutes. Interviews took place in-person in a private room during the study site visit, or online (using Zoom), using semi-structured interview guides adapted to the type of interview and provider (Supplementary file 1).

The online group discussions were organised with GPs attending continuous medical education sessions in local peer groups. We invited all peer groups in the Flanders region and held group discussions with those that agreed to participate. We conducted 16 group discussions (5-10 participants per group). All group discussions were conducted via Zoom, lasted between 80 and 100 minutes, and were moderated by two members of the research team: a qualitative researcher (JV) and a medical PrEP expert. We used an open-ended topic guide that included case vignettes and statements [40] to facilitate discussions on GPs’ involvement in PrEP care (supplementary file 2).

Lastly, we reviewed two policy documents to gain additional insight into macro-level factors governing the scope of practice around PrEP. These included: (1) the national PrEP reimbursement regulations [20]; and (2) the “HIV convention”, a funding contract between national reimbursement authorities and HIV clinics to finance their HIV and PrEP programmes [39].

### 2.3 Data analysis

Interviews and group discussions were audio-recorded and transcribed verbatim. Transcripts and documents were repeatedly read through and annotated by JV to support familiarisation with the entire dataset. During this phase, JV developed an initial set of codes, informed by close engagement with participants’ accounts and remaining open to concepts emerging from the data. These initial codes were partly inductive, while being situated within broader, theory-informed questions that guided the initial data exploration (Table 1).

**Table 1.** Theory-driven questions guiding initial data exploration.

| **Theoretical concept** | **Guiding question** |
| --- | --- |
| Professional identity | How do different types of providers make sense of the value they and their professional discipline add to quality PrEP care? |
| Boundary work | How do providers perceive their own (current or potential) role in PrEP care, and how do they relate to the role of others? |
| Role legitimacy | Who is perceived as a legitimate PrEP provider? |
| Competitive power | Where do providers leverage or express traces of competitive power? |
| Collaborative power | Where do providers leverage or express traces of collaborative power? |

Codes and emerging analytical narratives were discussed iteratively within the research team, resulting in successive adaptations of codes and refinement of the analytical focus. As analysis progressed, theoretical concepts of competitive and collaborative power were more explicitly mobilised to support interpretation, enabling systematic comparison across professionals and organisations. Throughout this process, coding involved moving iteratively between theoretical concepts, retrieved data segments and full transcripts (for context) to assess the fit of data with theory, explore variation, and consider alternative explanations. We used the qualitative data analysis software Nvivo (QSR; version 1.7.1) to support organisation, coding, and retrieval of data.

### 2.4 Reflexivity

The research team brought together interdisciplinary expertise: JV has a medical background with additional training in qualitative methods; GS has a background in psychology and mixed-methods sexual health research; TR is a sociologist with extensive experience in PrEP research; KK is a medical anthropologist with long-standing expertise in health systems research involving the health workforce; CN is a health psychologist with substantial experience in HIV research of policy and practice relevance. Ideas for this study emerged from JV’s doctoral research on PrEP service delivery in Belgium, and discussions within the Belgian PrEP Network Collaborative Care Working Group in which JV was previously a member and GS currently coordinates. All authors contributed to developing the theoretical framing, interpreting findings, and refining the analysis.

None of the authors were health workers. Although some authors were affiliated with a certified HIV clinic, this affiliation did not involve PrEP provision. The team’s shared interest in innovative and collaborative care models may have presented a source of analytical bias by orienting attention towards collaborative care and alternative configurations of PrEP delivery. We engaged regularly in reflexive discussion within the team, deliberately interrogating our own assumptions about professional collaboration and grounding interpretations closely in participants’ accounts, including those expressing scepticism towards alternative care models. The use of interdisciplinary perspectives and repeated discussion of emerging findings within and beyond the research team supported critical reflection on our positionality and helped mitigating favouring normative preferences over empirical findings.

### 2.5 Ethics statement

This research was approved by the Institutional Review Board of the Institute of Tropical Medicine (ref. 1416/20 and 1431/20) and the Ethics Committee of the University Hospital of Antwerp (ref. B3002020000223). Prior to participation, all participants provided informed consent; consent was audio-recorded (for interviewees) and written for participants in the online group discussions.

## 3. Results

We structured our findings according to three major themes, representing the distinct levels at which power dynamics and collaboration were enacted. We first expand on how formal policy and regulatory instruments influence power dynamics. Subsequently, we show how competitive and collaborative power shape the way boundary and collaborative work is enacted both among professionals and organisations in PrEP delivery.

### 3.1 Policy and regulation as structuring power dynamics

Professional jurisdiction over PrEP delivery was found to be formally consolidated through the HIV convention and PrEP reimbursement regulations, which respectively allocates funding and assigns PrEP initiation and prescribing authority to specialist physicians in HIV clinics. All providers, across professional cadres, described these documents as reference points, setting clear boundaries around who was authorised to act and with what degree of agency. As one specialist physician noted: “*For PrEP, we have to follow the convention. It defines who can prescribe and how follow-up should be organised*.” [HIV clinic 03]

Because funding and reporting obligations are tied to care delivered within HIV clinics, we identified few incentives to share PrEP activities with external providers. For GPs, the absence of prescribing authority and formal responsibility discouraged engagement, sending the message that PrEP “*is not really part of our job*” [GP213]. CBO representatives described tight PrEP regulations as prioritising institutional control at the expense of accessibility, particularly for marginalised populations.

While the HIV convention consolidated specialist jurisdiction, it was also reported to structure interprofessional collaboration within HIV clinics by mandating the involvement of other professionals such as nurses. Participants described how this enabled substantive task-sharing, including counselling and free on-site sexology consultations for PrEP users:

> *“The convention helped us to really kick-start the delegation of [PrEP] follow-up to our nurses […] this was really a necessity…our clinics were flooded with PrEP patients and […] I feel a bit overqualified to see them all three-monthly when there are no acute problems”* [HIV clinic 06]

While top-down issued PrEP regulations were clear, we found that the focus of PrEP care on prevention and risk management could cause ambiguity around professional jurisdiction. GPs and CBO representatives emphasised that PrEP did align with their prevention and care continuity ethos. Several HIV specialists also recognised that PrEP differed from HIV treatment in terms of specialisation, with some even stating that PrEP ultimately “*belongs*” in primary care. At the same time, these specialists emphasised that PrEP had become embedded in HIV clinics, with one specialist explicitly referencing competitive financing structures as a barrier to interorganisational collaboration:

> *“Ideally, [PrEP] is done completely in primary care […] But I think that the bottom line is also important, which people often don’t want to talk about, namely that we earn less money when we send all these patients to their GP. That’s of course no reason not to have GPs [involved in PrEP]. But unfortunately, it’s an element in the system that plays a role…”* [HIV clinic 01]

Nurses and sexologists in HIV clinics frequently added a layer to this ambiguity by framing PrEP as embedded in broader sexual health and psychosocial contexts that included complex interactions with issues of loneliness, transactional sexual encounters and substance use. Their accounts challenged a narrow biomedical framing of prevention and suggested that PrEP regulations should address unresolved questions on which health system actors ultimately carry responsibility over sexual health promotion and prevention.

### 3.2 Professional boundary work

When confronted with hypothetical scenarios of increasingly delegating PrEP care to GPs, HIV specialists often used claims of expertise and quality standards to legitimise their position as preferred PrEP providers. Many expressed concern over GPs’ capacity and competence in STI management and inclusive sexual health care. High levels of tacit knowledge, including on the needs of sexual minority patients, were deemed necessary to take on PrEP care, as reflected in this specialist’s comment on the complex needs of a ‘prototype’, high-risk PrEP candidate:

> *“I think we should defend our specificity and not separate PrEP from HIV [medicine], and recognise that when you see a lot of people [taking PrEP], your experience is making you better and better every day […] So I think it’s really, really important and I don’t think that if one [GPs] sees one MSM [man who has sex with men] from time to time, will you talk about drugs? Will you talk about other STIs, and HPV? Will you talk about all this stuff? I’m not sure”* [HIV clinic 06]

Nurses frequently nuanced these reflections by foregrounding relational dimensions of quality. Trust and a non-judgemental attitude were often described as foundational. They emphasised the need to adapt to patients’ concerns and experiences, describing their work as based on “*feeling*”. These accounts suggest that competitive power is not exercised solely through clinical-technical claims, but also through implicit hierarchies that privilege biomedical expertise over the emotional work of caring.

GPs and CBO representatives indirectly challenged a framing of quality grounded in expertise tied uniquely to sexual health. Whilst recognising their limited concrete experience with PrEP, GPs identified several areas where their specific expertise could be of added value. A GP reflects in the following quote on their advantage in being able to broaden access to PrEP through a holistic approach:

> *“We know people in their entirety, with their past and underlying issues […] I think we go about it more broadly, asking about your mental health status and why risk behaviour occurs in the first place.”* [GP322]

The asymmetry between strict and clear rules on paper, and underlying contestation and ambiguity in practice, highlights how competitive power operates through differing visions of legitimate standards of care rather than explicit conflict. Specialist physicians framed quality concerns through the lens of clinical responsibility and risk management that justified a degree of centralisation, whereas GPs, nurses and CBO representatives more frequently framed collaboration as inherently part of quality care.

Yet despite recognising its relevance, GPs rarely *claimed* jurisdiction over PrEP during group discussions. Most described limited engagement shaped by restricted authority, uncertainty and limited experience, which they attributed to regulatory restrictions and the cumulative effects of past organisational decisions. Several GPs noted that HIV had progressively disappeared from their daily work, reinforcing perceptions that PrEP initiation and follow-up required specialist knowledge, experience or infrastructure. Some comments reflected a sense of being on the periphery of HIV care:

> *“We [GPs] are deliberately bypassed by the system that keeps all this [HIV care] so centralised. So naturally we lose contact with it, as we don’t get the opportunity to build experience.”* [GP604]

Some GPs, particularly those younger or working in urban, multidisciplinary practices expressed an explicit desire to initiate PrEP without the need for referral to an HIV clinic. They framed PrEP as aligning with improving access for underserved groups that were part of their target population. Other GPs, often working in rural areas or monodisciplinary settings, described limited exposure and poor compensation of time-intensive activities such as counselling as challenges to integrate PrEP in routine practice. HIV specialists, however, often saw GPs as a homogeneous group insufficiently prepared or motivated to provide PrEP, implicitly contributing to the stabilisation of specialist jurisdiction. Overall, these accounts reveal subtle and uneven claims to professional authority that were shaped by historical factors, practice context, and commitment to equitable access.

Collaborative power, on the other hand, was more evident within HIV clinics through extensive task-sharing between physicians and nurses. Several nurses described having substantial autonomy within agreed protocols or informal arrangements that had grown organically, enabled by co-location, time, informal supervision, mutual trust and shared accountability under the HIV convention. Physicians described reliance on nursing expertise as essential for sustaining PrEP services at scale while not being perceived as threatening to medical jurisdiction, as direct supervision and prescribing authority remained within physicians’ control:

> *“They [nurses] help us to prioritise and take the load off […] we have trained them to see ‘OK these are normal blood values or symptoms, and this is suspicious’. When all the tests are negative, I totally trust them. If they are not sure, they come knocking on our door for advice. Like this we have a control over the ‘complicated PrEP’ in a way.”* [HIV clinic 06]

Nurses leveraged collaborative power through essential coordinating and relational work that structured everyday PrEP delivery. They described determining PrEP eligibility and organising personalised follow-up trajectories. Moreover, through their relational proximity with patients, nurses described being often more likely to attend to psychosocial concerns that may not surface in more hierarchical physician-led consultations. This relational positioning enabled nurses to act as internal boundary spanners by explicitly facilitating linkage to sexologists or psychologists. Collaborative power thus operated not only through task-sharing, but also through the creation of communicative spaces in which sensitive issues could be raised and discretionary decisions made about when and how to extend the internal care network.

Several nurses, however, also described taking on responsibilities that exceeded their formally defined scope. While nurses valued this autonomy and growing breadth of expertise, some expressed a desire for clearer recognition, training pathways, or formalisation:

> *“We now often work in ways that the law does not really cover […] I mean, the doctor writes the prescription because he is the one with the authority, but sometimes he knows less about the patient, so… Midwives in Belgium prescribe within their area of expertise, and I am sure there are lots of other places where nurses do it […] It would be good to officially have that independence through a qualification.”* [HIV clinic 08]

These accounts show that collaborative power within clinics reshaped, rather than eliminated, professional boundaries, shifting negotiation towards issues of recognition and long-term viability.

### 3.3 Organisational boundary work

Across organisational boundaries, competitive power became visible through examining PrEP care pathways. HIV clinics functioned as central entry points from where conditions for the involvement of other providers could be set. While specialists in one clinic reported systematically offering clients follow-up visits with their GP, most other HIV clinicians reported having only sporadic, patient-initiated, discussions on this topic. Where collaboration with GPs occurred, it was typically structured by specialists as informal, unilateral arrangements to address capacity constraints or in response to explicit patient requests, while key clinical decisions remained within HIV clinics’ control:

> *“We simply don’t have the capacity to see all these patients every 3 months […] So we started offering the option to have alternating visits with their GP […] We give them a prescription and a pre-filled laboratory request form to give to their GP, so they know what tests to perform.”* [HIV clinic 05]

Delegated GP roles were described as supportive rather than involving active decision-making, with overall responsibility for PrEP care remaining within HIV clinics. Some GPs voiced concerns over a mere tokenistic involvement and instead argued for true shared-care models:

> *“If we are to be more involved in [PrEP care], we must make sure that our role does not become a formality or a convenient resource for specialists. We have had a nine-year medical training, and collaboration should happen on equal footing.”* [GP532]

GPs described inconsistent communication from HIV clinics, particularly when patients requested PrEP directly in the HIV clinic. This could result in role blurring when patients later came to their GP for follow-up:

> *“I find it difficult that you don’t really know what is going on in the [HIV clinic]. When patients come to you to request a [PrEP] refill, I don’t know to what to do. Do I still need to counsel? Do I need to explain anything? Has there been adequate screening? You don’t know where your role starts or ends.”* [GP005]

HIV specialists acknowledged these challenges and described communication with GPs as difficult to address within existing organisational structures and privileged patient-provider relationships. As a result, collaborative practices often remained *ad-hoc* and dependent on interprofessional relationships rather than shared systems:

> *“It would be better to send a written report of our consultation to GPs, but I think some patients will not be happy […] They like that their GP does not have to know about their sexual life and behaviour.”* [HIV clinic 03]

Nurses in one HIV clinic that implemented a more systematic, informal task-shifting of PrEP follow-up practices to GPs, noted that effective collaboration required active contact and shared understanding. They mentioned being frequently confronted with calls from GPs reflecting hesitancy and uncertainty on their role in PrEP delivery, which prompted the organisation of a webinar on PrEP to address these issues. Their accounts indicate that interorganisational boundary work needs to be mediated through mutual buy-in and systematic communication.

Unlike GPs, CBOs described HIV prevention as core to their historical and organisational mandate. Several CBO representatives, especially those working with undocumented sex workers excluded from formal PrEP eligibility criteria, described facilitating or directly providing PrEP through medical doctors embedded within their organisations. HIV specialists recognised this and enabled their practice through collaboratively developing informal, locally negotiated protocols. While these protocols reflected collaborative power between HIV clinics and CBOs, they also functioned as mechanisms that allowed HIV clinics to maintain a degree of oversight.

We found that CBOs had a crucial role connecting individuals to existing services and compensating for accessibility barriers created by centralised arrangements. CBO representatives described actively facilitating access by arranging appointments in real-time and accompanying patients to the HIV clinic:

> *“What I tell clients coming to request PrEP is ‘okay you want to start PrEP, we are going to start today. And the first step is that we talk about PrEP, do your testing and then you go to a [HIV clinic]’. And I call the clinic with the phone on speaker and say ‘I have this patient who wants to start PrEP […] When could you see him, when do you have an appointment?’ This makes people feel like there is a partnership and that they have started PrEP already.”* [CBO representative 02]

CBO representatives also described adapting to regulatory constraints, particularly by negotiating pragmatic solutions for uninsured or undocumented clients, leveraging personal relationships with specific clinicians:

> *“I know which doctor is flexible on the conditions about PrEP [initiation] and which doctor is stricter. So, I try to have my favourite partners for different things. Like, I know some doctors will give PrEP from their own informal stock and others won’t do it.”* [CBO representative 01]

In doing so, CBOs did not challenge specialist control over care pathways but exercised collaborative power by bridging organisational boundaries and prioritising access for marginalised populations.

Taken together, we showed that PrEP scale-up in Belgium is shaped by power dynamics across professional and organisational boundaries, as summarised in Figure 1. Professional boundaries, roles and tasks in PrEP care are being set and protected (i.e. competitive power) using regulation that currently favours specialist jurisdiction over PrEP delivery. Collaborative power develops where organisational arrangements support interdependence, as observed within HIV clinics through nurse-physician task-sharing and a multidisciplinary team-based approach. Across organisations, collaboration remained rather informal and relationship-based.

**Figure 1.**
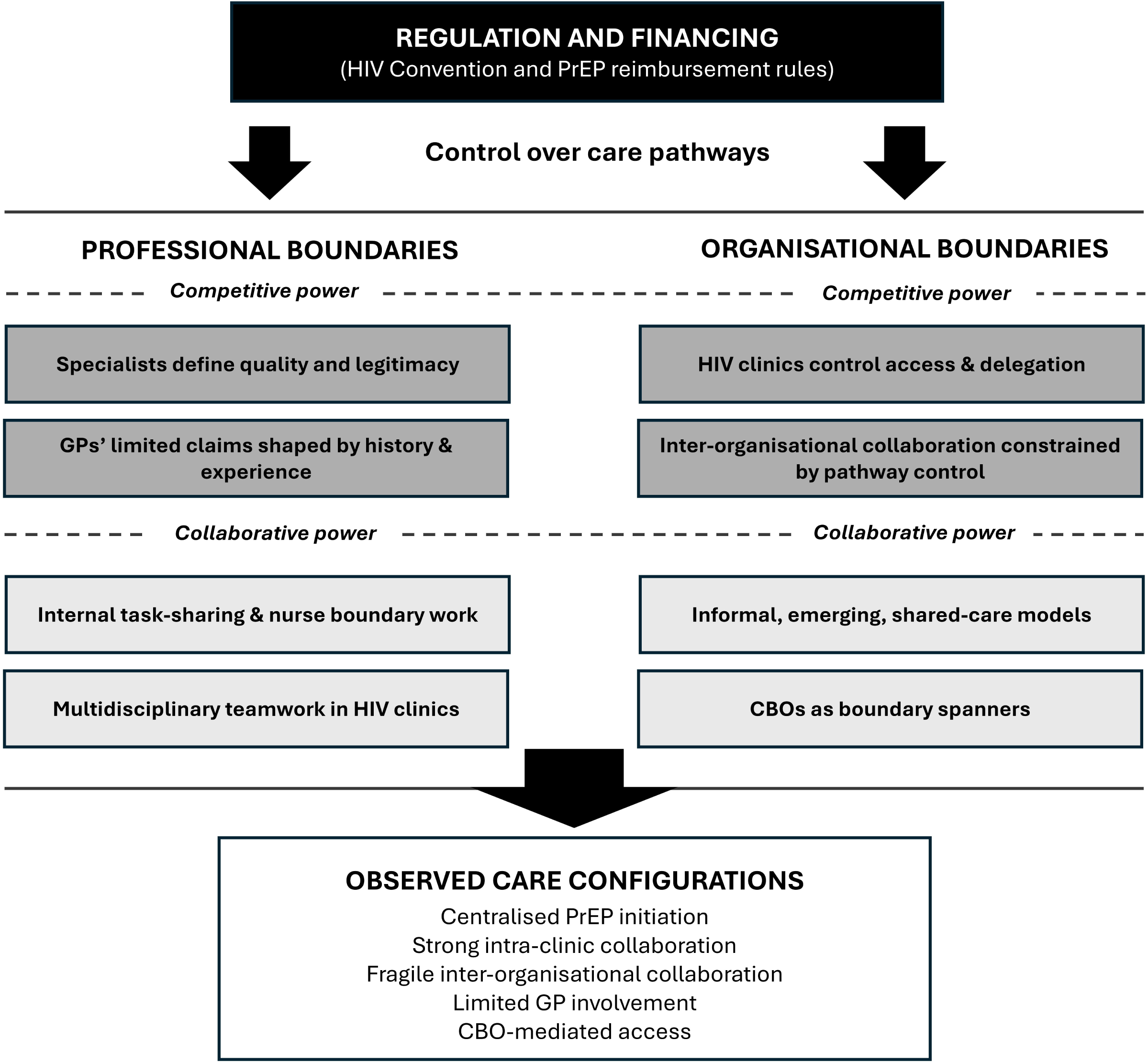
Formal rules and regulations influence how competitive and collaborative power operate to shape PrEP care configurations across professional and organisational boundaries in Belgium.

## 4. Discussion

Our study applied a theoretical lens on power and collaboration to better understand how providers negotiated expertise and role division to expand PrEP care beyond specialised HIV clinics in Belgium. Participants consistently linked professional roles in PrEP care to structural, regulatory and funding arrangements, indicating that observed professional tensions are less the result of individual resistance than of institutional arrangements that allocate authority, resources and accountability unevenly across health system actors. These findings help to explain why collaborative PrEP delivery often remains partial despite broad rhetorical support for integration.

This research advances theory by offering insights into the influence of power beyond teams, and across health system levels. While competitive and collaborative power co-exist, the conditions enabling collaborative power at one level may be absent at another. Within HIV clinics, interprofessional collaboration was facilitated by shared organisational parameters (e.g. co-location and low-threshold communication), formal reimbursement mechanisms, and clear accountability structures. This is consistent with existing literature on interprofessional collaboration; where professionals work within the same organisation, share the same vision and goals, and operate under mutually agreed protocols, mutual trust can develop and collaboration becomes routinised without fundamentally challenging established hierarchies [33,41].

At the interorganisational level, structurally enabling conditions were largely absent in our study. Collaboration between HIV clinics and GP practices was facilitated *ad-hoc* and informally, often driven by capacity constraints in HIV clinics rather than shared governance arrangements. This aligns with existing evidence showing that interorganisational collaboration is typically more difficult to achieve than interprofessional collaboration, as it requires alignment not only of professional identities, but also of organisational contexts, funding mechanisms and accountability systems [33,42]. In our study, the absence of shared communication systems and formal coordination mechanisms, persistent role blurring, and an historically embedded lack of confidence in the HIV expertise of GPs, limited the leverage for collaborative power across organisational boundaries.

Our work shows that, in regulatory environments where boundaries are formally clarified, boundary work shifts from open professional conflict to more subtle forms of contestation. This became visible in narratives about expertise and quality, where specialist care emerged as a reference point for maintaining clinical standards while participants raised genuine concerns about how quality could be ensured across more distributed delivery models. While safeguarding clinical quality and safety is essential, equating quality primarily with specialised delivery risks overlooking important systemic dimensions of access and person-centred care, and creating a space where expertise is privileged rather than shared among health system actors [43]. Importantly, recent evidence suggests that not all PrEP users require the same intensity of follow-up nor the same package of care at each visit, suggesting PrEP service delivery should allow for sufficient differentiation according to user needs and preferences [44–46]. A 2022 study among a non-random sample of 326 Belgian PrEP users showed that 63.8% preferred the HIV clinic for follow-up, while 51.9% were willing to have their GP involved, signalling considerable patient support for collaborative care models [47]. From a public health perspective, centralising service delivery may undermine impact at the system level by limiting access. This issue is of relevance in Belgium, where a considerable group of people at risk of HIV remains underserved (e.g. clients with intersecting vulnerabilities, immigrant men having sex with men, and women of sub-Saharan African descent) [48,49].

### 4.1 Implications for collaborative PrEP care models

The global health literature consistently shows that nurse-led and primary care-based PrEP provision is safe, effective, and cost-efficient [50–52]. However, implementation at scale often remains limited, even in well-resourced settings with near-universal health coverage, such as Belgium. Our study underscores that barriers to delegation and collaboration in PrEP delivery are not just clinical or technical (e.g. resource-related), but also fundamentally structural and political. Consequently, interventions focused solely on training, guidelines or evidence dissemination are unlikely to succeed if they do not address underlying power relations and organisational incentives. This is a vastly under-researched domain in the PrEP literature with important practical and policy implications given ongoing international debates on how PrEP care should be organised in, for instance, the Netherlands [53], Australia [11,46], South Africa [54] and the United States of America [55]. For instance, the South African Medical Association legally opposed the Pharmacist-Initiated Management of Antiretroviral Therapy programme, arguing that pharmacists lack essential training and skills to initiate PrEP [56]. Despite this opposition, the High Court in 2023 ruled that trained pharmacists are allowed to prescribe PrEP, though the programme continues to face appeals from physicians [57]. Our research offers a framework to start interrogating the role of power dynamics, jurisdiction and collaboration in PrEP, an issue of continued importance with the arrival of novel PrEP formulations involving new sets of (clinical) practices (e.g. injectables and implants) [58].

Secondly, our findings focusing on the Belgian situation suggest that achieving professional engagement and collaboration in PrEP requires looking beyond individual attitudes (i.e. as phrased in the “*purview paradox*” [10]). Existing research from chronic disease management and mental health shows that collaborative care can be cost-effective, team-based models to support population-level and patient-centred outcomes, provided that formal integration mechanisms are present: shared protocols, designated coordinators, systematic communication channels, and bundled funding [59–62]. Such mechanisms are essential for scaling collaboration beyond pilot sites and motivated individuals. Our study findings suggest that similar principles apply to PrEP care but are not yet fully realised in health systems, including in Belgium.

Thirdly, our findings underscore the need to bring CBOs more explicitly into collaborative care studies and models. Literature on pluralistic health systems shows that non-state actors often play crucial, yet under-recognised (including unremunerated) roles in bridging access gaps and building trust [35,63]. Their crucial role in reducing PrEP-related disparities is, however, increasingly recognised, with innovative community-based PrEP delivery models emerging across Europe [64]. Our data suggest that effective PrEP scale-up requires clearly defined, accountable partnerships between CBOs and formal health services. Future research is needed that contributes to a clear understanding of the roles, accountabilities, and integration of CBO roles with more conventional health system actors (e.g. clinicians) and systems (e.g. funding).

### 4.2 Policy and practice recommendations

While Belgian PrEP regulations sought to ensure quality through specialist provision, the concentration of authority and resources within HIV clinics generated capacity bottlenecks and constraints for interorganisational collaboration. Recent incremental reforms tolerating limited GP involvement do not fundamentally alter these dynamics, illustrating how piecemeal adjustments are likely to fail as long as macro-level regulations, meso-level organisational incentives, and micro-level professional interests remain misaligned. Transforming these patterns demands structural reforms: regulatory changes that genuinely enable shared responsibility and limit role ambiguity, formalising and recognising the expanded scope of nursing practice and expertise in PrEP, funding mechanisms that reward coordinated care and preventive work in primary care, and formal infrastructure that supports systematic rather than *ad-hoc* communication and collaboration.

Research on clinical networks and communities of practice shows that interdisciplinary platforms can strengthen collaboration by building trust, shared norms and values, and collective problem-solving across organisational boundaries [41,65,66]. Regular interaction around shared tasks improves relational coordination and mutual understanding, which helps overcome professional silos and contributes to the “software” needed for successful collaboration [67]. In this light, the Belgian PrEP Network – established in 2021 as an informal platform for regular exchange on the Belgian PrEP roll-out among HIV clinicians, GPs, researchers, CBOs and other relevant public health actors – has fostered an informal community of practice that supported shared-care development within existing constraints [68]. Policymakers should invest in nurturing these collective spaces and leverage them as governance instruments that can help build capacity for PrEP care, align quality standards, and enable scale-up.

### 4.3 Study limitations

This study was conducted within a specific context, and findings may not be directly applicable to settings with different health system parameters. However, through rich description and explicit theory engagement, we have attempted to produce transferable insights. Secondly, our participants consisted predominantly of HIV specialists, nurses and GPs. We only included CBO representatives who were reported by HIV clinics as existing collaboration partners. Other providers with stakes in PrEP care (e.g. other CBOs, pharmacists, social workers) were not included but likely have relevant insights to contribute. The policy landscape in PrEP service delivery evolves rapidly (e.g. data collection preceded the 2023 regulatory revision tolerating limited GP involvement), underscoring the need for continued empirical monitoring of how new mandates reshape collaboration arrangements and access to care over time.

## 5. Conclusion

Professional collaboration in providing PrEP care is shaped by power relations that operate differently across professional and organisational boundaries. We found that interprofessional collaboration within HIV clinics was facilitated by supportive structural conditions, while interorganisational collaboration remained constrained by regulatory, organisational and historical factors. By highlighting how collaborative and competitive power co-exist and are enabled by different conditions, this study contributes to a theory-driven and nuanced understanding of how PrEP care can be organised. Strengthening integrated care and prevention models will require not only training and willingness, but sustained investment in governance, funding, and shared platforms that enable trust and coordination.

## Data Availability

The entire qualitative datasets presented in this article are not readily publicly available because they contain information that could compromise the privacy of our research participants. Additional data are available from the first author on reasonable request.

## Notes

### Competing Interest Statement

The authors have declared no competing interest.

### Author Declarations

This research was approved by the Institutional Review Board of the Institute of Tropical Medicine (ref. 1416/20 and 1431/20) and the Ethics Committee of the University Hospital of Antwerp (ref. B3002020000223).

### Funding

- Research Foundation - Flanders: S004919N

