## Supplementary File 1 for "Re-shaping professional boundaries to scale-up HIV pre-exposure prophylaxis (PrEP) services: collaborative care and power dynamics in Belgium"

**Semi-structured interview guide for health care providers in HIV clinics.**

1. **<u>Respondent background</u>**

- ***Can you tell me something about yourself in terms of educational background, professional career and current activities you perform as part of your job?***
- ***How would you describe your role in the HIV Reference Center? And can you tell me some more about the work you do around PrEP?***

2. **<u>The PrEP care process and notions of required skills and expertise</u>**

*PrEP is a relatively new intervention, and we are still learning how to best provide it. I would like to take some time to go through your experience with providing care for PrEP clients, and your thoughts on how care is organized here at the HRC. Is that okay with you? [confirmation from participant] Great, so let me start with getting a bit of a clearer picture of what happens exactly at a PrEP consultation here at the HRC.*

- ***Imagine that I am a first-time client coming to your consultation for PrEP, can you walk me through what would happen? How would you usually handle such a first-time visit?***
    - When would I first come into contact with you?
    - Where are you located? (e.g. separate cabinet or desk? Proximity to other providers?)
    - What do you do during a typical PrEP visit (focus on first-time visit)?
        - Eligibility screening
        - Education
        - Counseling
        - Lab testing
        - PrEP prescription
        - Vaccinations
    - Which other providers would I meet before or after visiting you? What would they still do on top of what you do?
    - How is this different or similar from follow-up visits (e.g. would I meet you again every time?)

- ***You mentioned to me different care aspects that are part of a typical PrEP visit. Let me go over each and one of them, and get some more insight of what they all require of you.***
    - *<u>Assessing who are appropriate PrEP candidates</u>*: how do you do this? Are there certain criteria you apply? Do you follow the reimbursement criteria? Why (not)?
    - *<u>Providing education on PrEP</u>* : Dou you have a standard way of going about this? What skills would you say it takes to provide good education on PrEP? What helps you to provide

good information (e.g. are there any materials you use to support your message?). Would you tailor this information according to the type of client in front of you? In what way?

- *Providing counseling on sexual health*: Apart from PrEP as such, do you engage in a conversation on sexual health in general (e.g. risk of other STIs, condom use, other prevention available options, family planning etc.)? Why (not)? Some providers might feel uncomfortable talking about sex, can you relate to that? Are there any materials you use to support your message?

- *Prescribing laboratory tests*: Who decides which laboratory tests are done at a PrEP visit? What is this based on (e.g. guidelines, client profile or history, consensus in the team etc.)? Who can prescribe the necessary laboratory testing?

- *Prescribing PrEP*: Who delivers the prescription for PrEP? How much PrEP is prescribed at each time (e.g. multi-month dispensing)? Does this sometimes vary? If yes, when? Who decides this?

- *Providing follow-up care:* What does follow-up for PrEP clients entail once they have started on a PrEP regimen? How are you involved in this? Does the frequency and nature of follow-up sometimes vary between individuals? Can you give some examples of how and when it can be different?

- *Responding to other sexual an mental health needs*: Are you sometimes confronted with other sexual and mental health needs of clients? How do you deal with this? What skills or expertise do you require to meet needs and demands from clients in this regard?

- ***As you might have noticed, I am interested in trying to describe what it takes to "do PrEP well", in terms of skills, expertise, values and practices. Can you name three things (e.g. specific skills, attitudes or values) that, in your experience, are vital to ensure 'good quality care' for PrEP?***
  - Why the choice for these three elements? How would you rank them according to importance
  - How do you implement these three things in your day-to-day practice?
  - What has supported you, or still supports you, to provide good quality care for PrEP clients?
  - What barriers do you sometimes encounter to live up to putting these three things into practice? Can you give examples?
  - What does it take for one to acquire this expertise?
  - How does this expertise relate to the following factors (*probe for each*):
    - professional training
    - personal norms and values
    - being part of a multidisciplinary team
    - access to certain resources (e.g. equipment, guidelines or infrastructure)
    - experience (years in the field ; engaging regularly in sexual health care)

- being part of a key population for PrEP/knowing the key populations well
- other?

### 3. Client-centered care and shared decision making

*In the provision of health care, the concept of "patient-centered" or "client-centered" services has been receiving more and more attention. This approach means "providing care that is respectful of, and responsive to, individual patient preferences, needs and values, and ensuring that patient values guide all clinical decisions".*

- ***When you think again about – let's say – the last five to ten clients you attended to for PrEP, how would you say you were able to implement 'client-centered care' ?***
    - (*to stimulate thinking*) Would you say you treat everybody the same?
    - How were you able to identify and respond to specific clients' needs?
    - How were you able to identify and respond to clients' specific preferences?

- ***One aspect of "client-centered care" is shared decision making between the provider and client. To what extent do you engage clients themselves in decisions for each of the following care aspects:***
    - *Whether or not PrEP is a good option for clients*: How do you support clients in this decision? Would you sometimes advise against PrEP? When and why? Would you sometimes advise alternative HIV prevention options instead of PrEP? When and why?
    - *Which PrEP regimen to start (daily or event-driven)*: How do you support clients in this decision? Would you sometimes advise against certain regimens? When and why?
    - *Which laboratory tests to perform*: To what extent can clients choose which tests are performed as part of screening or follow-up (incl. biochemistry, organ functions, HIV and STI testing)? Which tests can be more flexible than others? Are the same tests always performed in every individual? Why (not)?
    - *Which vaccines to get (incl. HPV)*: How are vaccinations discussed with clients? Would you recommend an HPV vaccine to all individuals? Why (not)?
    - *Plans around the practice of 'safe sex' (e.g. condom use)*: Do what extent is a strategy for protection of other STIs (besides HIV) being discussed with all clients? How would you describe your responsibility versus the responsibility of clients when it comes to sexual health promotion and prevention?
    - *On timing (date and time) of follow-up visits (for testing and consultation)*: To what extent do clients have a choice when to return for a follow-up visit (frequency and timing during the week) ? Can clients visit the same provider every time? How important would you think it is to have the same provider attending to clients?

- *On how to involve clients' GP in PrEP care*: To what extent do you discuss with every new client how they would like their GP to be involved in PrEP care? If yes, why is it important to do this? If not, why is it not always discussed?

- ***In your experience, what can be barriers to practice 'shared decision making' in PrEP care.***
    - When is it not desirable? Why?
    - When is it not feasible? Why?
    - Which factors can make it difficult to practice 'shared decision making'
        - probe for policy, health systems, facility-related and provider-related factors

- ***In your experience, what are crucial facilitators to practice 'shared decision making' in PrEP care?***
    - Which practices or interventions stimulate clients' engagement?
    - How can the working of the HRC be made more conducive to practicing 'shared decision making' ?

**4. Reflections on the service delivery model for PrEP**

- ***Based on your experience, what would you say are three key advantages of providing PrEP through a specialized setting such as the HRC? Why do you think that is?***
    - What are some disadvantages of providing PrEP through the HRC? Why do you think that?
    - Around the world, people are thinking of what good service delivery models for PrEP could look like. There is increasing attention for the role of nurses on the one hand, and first-line health workers, such as family physicians, on the other hand. From your experience, what could be the added value of involving clients' family physician in PrEP care?
    - Alternatively, according to you, what conditions would be required for family physicians to be more involved in the delivery of PrEP? Why is that?
    - Do you feel clients could benefit of more collaboration between the HRC and their family physician? If yes, how could this collaboration look like? If no, why is that?
    - Another strategy I have observed in different HRCs, is task-shifting and task-sharing with nurses. In what way does this strategy benefit clients? And in what way does it benefit the providers at the HRC?

- ***To what extent do you feel that PrEP services at the HRC are a good fit for all clients who could benefit of its use? Why is that so?***
    - Differentiate between:

        - Geographical access (distance to the facility)
        - Acceptability (e.g. opening hours, waiting times)
        - Affordability (cost & reimbursement regulations)
        - Accommodation (how setting is constructed, “LGBTQ-friendliness”, ensuring a ‘safe’ and inclusive environment)
        - Availability (sufficient staff and resources to attend timely to clients)
    - Are there any groups that are insufficiently reached by the PrEP services at the HRC? If yes, which ones and why do you think that is?
    - In your opinion, what could be done to reach those groups better? For instance, are there other providers outside the HRC that could be involved?
