## Supplementary File 2 for "Re-shaping professional boundaries to scale-up HIV pre-exposure prophylaxis (PrEP) services: collaborative care and power dynamics in Belgium"

**Topic guide for online group discussions with general practitioners.**

| Action | Topic | Question/statement |
|---|---|---|
| Opening | Acquaintances | *Can you tell us who you are, where you work, and one thing you like about being a family physician (FP)?* |
| Introduction to topic | Perceptions and experiences of sexual health | *What activities and/or situations are you confronted with in your practice when it comes to sexual health?* |
| Case | Introduce case vignette #1 | |
| Key question | Talking about sex in FP practice | *How frequently do you discuss issues of sexuality with clients?* |
| | | *What are concrete entry points to discuss sexual health with clients?* |
| Case | Continuation vignette #1 | |
| Key question | Role of the FP in sexual health | *Statement: "It is the role of the FP to guide clients in the accomplishment of sexually healthy lives". (agree or not agree) + explain* |
| Case | Explanation on general aspects of PrEP | |
| Key question | Role of the FP in identifying PrEP candidates | *Statement: "It is the role of the FP to identify clients who could benefit of PrEP". (agree or not agree) + explain* |
| Case | Explanation on clinical aspects of initiating clients on PrEP | |
| Key question | Role of the FP in starting clients on PrEP | *Statement: "Initiating clients on a PrEP regimen can also fall under the responsibilities of the FP". (agree or not agree) + explain* |

| | | |
|---|---|---|
| Case | Introducing case vignette #2 | |
| Key question | Role of the FP in follow-up of PrEP users | *Statement: "Follow-up of clients on PrEP can be part of the responsibilities of the FP". (agree or not agree) + explain* |
| Case | Explanation on clinical aspects of PrEP follow-up | |
| Key question | Collaboration with specialist physicians | *What could a good collaboration between specialists and FPs for PrEP look like?* |
| Closing | Wrap-up | *Short summary of main discussion points and room for additional comments and/or suggestions from participants.* |
